# Patterns and Changes in Expectation of Life at Birth in India: 1998-2017

**DOI:** 10.1101/2021.04.15.21255592

**Authors:** Aalok Ranjan Chaurasia

## Abstract

This paper analyses patterns and changes in the expectation of life at birth in India and decomposes the increase in the expectation of life at birth between 1998-2002 and 2013-2017. The analysis reveals considerable volatility in the increase in the expectation of life at birth in the country and in its different population groups and states. In recent years, there is considerable deceleration in the increase in the expectation of life at birth in the country because of the deceleration in the increase in female expectation of life at birth. The decomposition exercise reveals that most of the increase in the expectation of life at birth is attributed to the improvement in the survival probability in the first five years of life. The analysis also suggests that the recent deceleration in the increase in female expectation of life at birth may be attributed to the decrease in the person-years lived in the age group 40-65 years.

## 1. Introduction

The abridged life tables prepared by the Registrar General and Census Commissioner of India suggest that the expectation of life at birth (*e*_0_) in India increased by more than 6 years from 62.9 years during 1998-2002 to around 69 years during 2013-17 (Government of India, 2019). The increase has been more rapid in females (6.4 years) than in males (5.9 years). When compared with the United Nations model mortality improvement schedules (United Nations 2004), male mortality improvement in India has been somewhere between slow to medium model mortality improvement trajectory while female mortality improvement has been somewhere between medium to fast model mortality improvement trajectory. According to the estimates prepared by the United Nations Population Division, the *e*_0_ in India was around 69 years during 2015-2020 which is 3 years lower than the world average of around 72 years (United Nations, 2019). Estimates prepared by United Nations Population Division also suggest that India ranks 144 among 201 countries in terms of *e*_0_ which suggests that the expectation of life at birth in India is low by international standards.

India was one of the signatories of the Programme of Action adopted at the 1994 International Conference on Population and Development at Cairo (UNFPA, 2004). One of the goals of the Programme of Action was that every country would take appropriate steps to increase *e*_0_ to more than 70 years by the year 2005 and to more than 75 years by the year 2015. Viewed from this perspective, the improvement in the expectation of life at birth in India has fallen substantially short of what was committed way back in 1994. Interestingly, India’s latest National Health Policy 2017, now aims at increasing the *e*_0_ to 70 years by the year 2025 (Government of India, 2017) which implies a significant lowering of the target of improvement in *e*_0_ as compared to what was envisaged and agreed upon by India more than 25 years ago.

Within the country, there are marked differentials and regional variations in *e*_0_. The *e*_0_ is higher in females compared to males and in urban areas compared to rural areas of the country. Among 17 major states of the country – states with a population of at least 10 million – for which official abridged life tables are available, *e*_0_ ranged from more than 75 years in Kerala to 65 years in Uttar Pradesh during 2013-17. Estimates of *e*_0_ for smaller states and Union Territories are not available. There are only six states, in addition to Kerala, where *e*_0_ is estimated to be more than 70 years during the period 2013-17. The female-male gap in *e*_0_ has increased from around 2.1 years in 1998-2002 to 2.6 years in 2013-2017 but the urban-rural gap has decreased from 6 years to 4.7 years during this period. Similarly, the gap between the highest and the lowest *e*_0_ across 17 states has narrowed down from around 13.9 years during 1998-2002 to around 10.2 years during 2013-2017.

**Figure 1:**
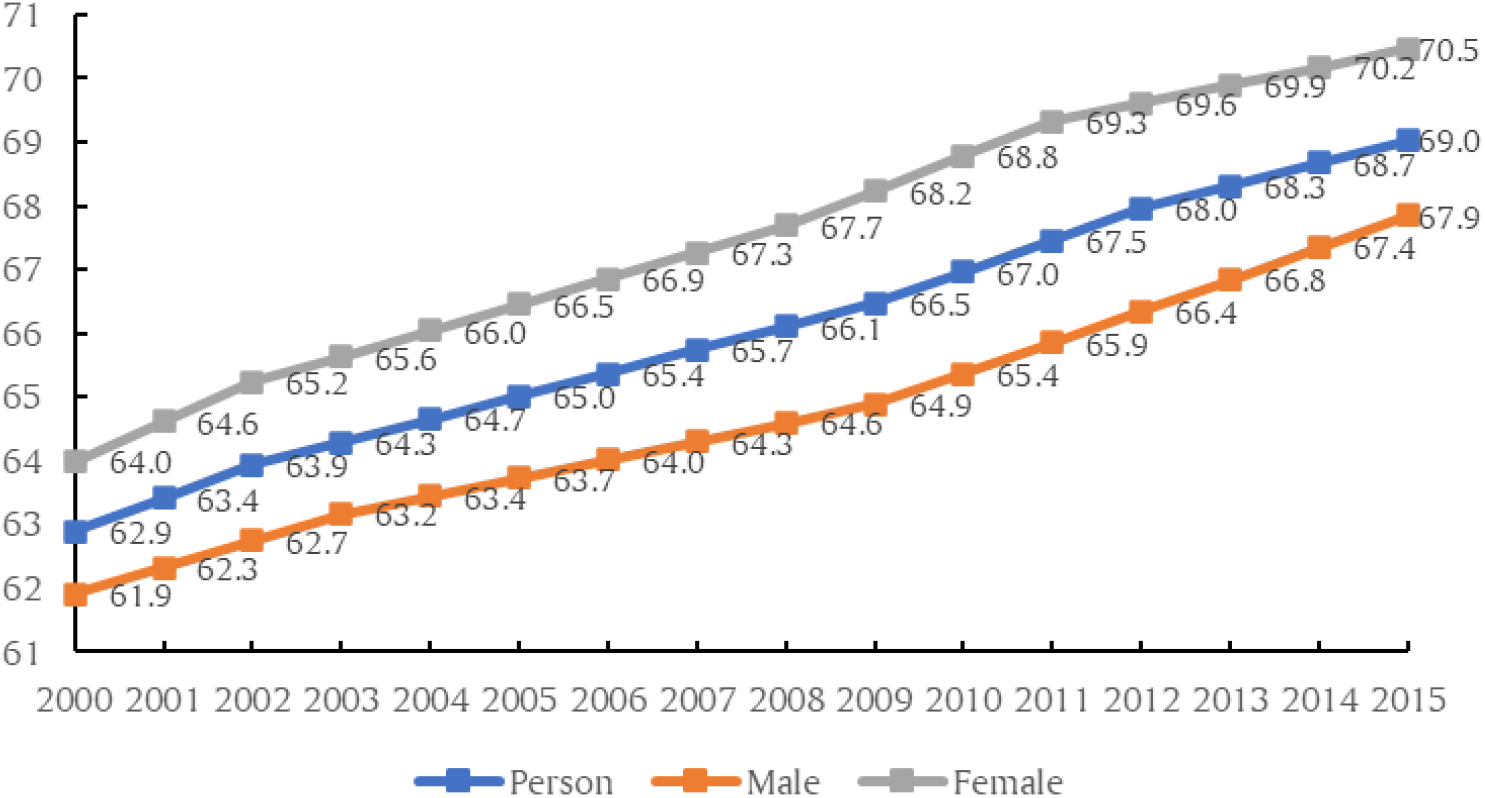
Trend in expectation of life at birth (years) in India, 2000-2015.

The expectation of life at birth is the most widely used summary indicator of population health (Wilmoth, 2000) – the greater the *e*_0_ the better the assumed health status of the people. Viewed from this perspective, the low level of *e*_0_ reflects the unsatisfactory state of population health in India by international standards. The expectation of life at birth, essentially, summarises the survival experience of the entire population. The relationship between survival experience of the population and *e*_0_, although reciprocal, is, however, complicated (Pollard, 1982). The impact of the improvement in survival probability at different ages of the life on the improvement in *e*_0_ is different. Monitoring changes in *e*_0_, however, is relevant as the increase in the length of life has always been one of the key health and development agenda throughout the world. Improvement in the health status of the people and reduction in mortality are widely recognised as the most proximate approaches of increasing the length of the life.

Despite the slow transition and marked within country variations in *e*_0_ in India, there is virtually no study, to the best of our knowledge, that has analysed temporal patterns, differentials and regional variations in *e*_0_ in the country in recent years. There have been many studies in the past that have analysed mortality transition in the country (Chaurasia, 2010; Mari Bhat, 1987) but recent studies on spatio-temporal variations in *e*_0_, especially, after 2000, are rare. Such an analysis appears timely as major policy level changes have been introduced in the country after 2000 that may be conjectured to have an impact on longevity. India announced a new population policy in 2000 (Government of India, 2000), followed by a new health policy in 2002 (Government of India, 2002). The National Rural Health Mission was launched in the year 2005 with a focus on establishing a fully functional, community-owned, decentralized health care delivery system (Government of India, 2005). In 2013, the National Urban Health Mission was launched (Government of India, 2013) and the two Missions were clubbed into National Health Mission in 2013 which envisages achievement of universal access to equitable, affordable and quality health care services that are accountable and responsive to health and family welfare needs of the people (Government of India, 2013). India has also recorded an unprecedented economic growth during this period. The country recorded an average annual growth rate of almost 7.7 per cent per year in the gross domestic product during 2001-2011 (Government of India, 2018). Although, economic growth slowed down after 2011, yet, it remained amongst the highest in the world. It is, however, not known how population and health related policy measures and rapid economic growth during have contributed to improvement in population health in India. The present paper is an exposition in this direction.

The paper is organised as follows. The next section describes the data source while the third section outlines the methodology. Trend in *e*_0_ in India are analysed in section four of the paper. Section five decomposes the increase in e0 into the change in survival experience in different age groups. Section six analyses regional variations in the increase in *e*_0_. The last section summarises the findings of the analysis from the perspective of population health in India.

## 2. Data Source

The analysis is based on the abridged life tables prepared by the Registrar General and Census Commissioner of India on the basis of the age-specific mortality rates generated through India’s official sample registration system (SRS). SRS is a large-scale demographic sample survey based on the mechanism of a dual record system which was launched in 1964-65 on a pilot basis to provide reliable estimates of fertility and mortality indicators. Since 1969-70, SRS covers the entire country (Government of India, 1971). Reporting of births and deaths in SRS has been found to be fairly reliable, although, there is some under-reporting of vital events under the system (Government of India, 1983; Government of India, 1988; Mari Bhat, 2002; Mahapatra, 2010; 2017). These abridged life tables are available for the country and for states having least 10 million population. Five years average age-specific death rates available through SRS are used for the construction of life tables to adjust for sampling fluctuations and to augment the sample size (Government of India, 2019). These life tables, therefore, reflect the average mortality experience of the population over a period of five years and it is assumed for the present analysis that the five-year average mortality experience actually refers to the mid-year of the five-year interval. Thus, abridged life table for the period 1998-2002 is assumed to reflect the mortality situation that has prevailed around the year 2000. In situation where no death is reported under SRS in an age-group, the age-specific death rate for that age group has been imputed on the basis of a geographic approach (Government of India, 2019). The abridged life tables so prepared are available for concurrent five-year periods since 1986-90 (1988). They are the only source to analyse temporal patterns of e0 in India. The present analysis uses abridged life table since 1998-2002.

## 3. Methods

The analysis has been carried out in two parts. The first part is devoted to analysing the trend in *e*_0_ while the second part analyses the contribution of the change in age-specific survival probabilities to the change in *e*_0_. The trend analysis has been carried out following the assumption that the trend in *e*_0_ may be influenced by policies and programmes directed towards increasing improving the health of the people and by improvements in the standard of living. This means that the trend in *e*_0_ may not be linear (on a log scale) but may have changed during the period under reference. Once the point(s) in time when the trend in *e*_0_ has changed are identified, the analysis is carried out separately for temporal segments between two points assuming that the trend is linear (on a log scale) in a given temporal segment.

There are different methods for identifying the time of change in trend including permutation test (Kim et al, (2000), Bayesian Information Criterion (BIC) (Kim et al, 2009), BIC3 (Kim and Kim (2016) and Modified BIC (Zhang and Siegmund (2007). The permutation test is the gold standard but is computationally very intensive. BIC performs well to detect a change with a small effect size but has a tendency of over-estimating number of points when the trend has changed. Modified BIC is the most conservative but performs well to detect a change with a large effect size. Performance of BIC3 is comparable to that of the permutation test.

Assuming that the trend is not linear (on a log scale), the trend analysis may be carried out through joinpoint regression analysis. The difference between the joinpoint regression analysis the conventional piecewise regression analysis is that the time or joinpoints when the trend has changed are determined from the data in joinpoint regression whereas they are decided or fixed beforehand in the conventional piecewise regression. Let *y*_i_ denotes the expectation of life at birth (*e*_0_) for year *t*_i_ such that *t*_1_<*t*_2_<…<*t*_n_. Then the joinpoint regression model is defined as

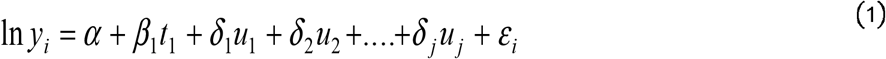

where

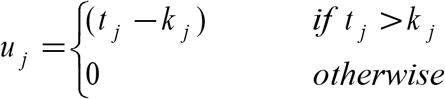

and *k*_1_<*k*_2_….…<*k*_j_ are the years when the trend has changed or joinpoints. Details of joinpoint regression are given elsewhere (Kim et al, 2000; Kim et al, 2004). Assuming that the trend is linear (on a log scale) in a temporal segment or between two joinpoints or

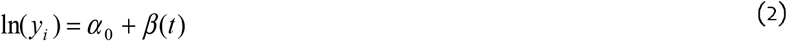

then the annual per cent change (APC) between two joinpoints or in a temporal segment is estimated as

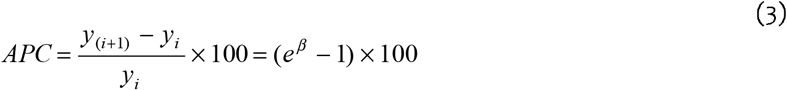

The average annual per cent change (AAPC) during the entire reference period is then obtained as the weighted average of APCs in different temporal segments with weights equal to the length of different temporal segments. The AAPC is argued to be a better approach to describe the long-term trend when the trend changes over time in comparison to the commonly used approach in which a single regression line on a log scale is fitted for the entire reference period and the average annual per cent change is calculated from the slope of the regression line (Clegg et al, 2009). AAPC permits comparison of trend in different temporal segments.

Actual calculations were carried out using the Joinpoint Regression Program (National Institute of Cancer, 2013). The software requires specification of minimum (0) and maximum number of joinpoints (>0) up to a maximum of 9 joinpoints in advance. The programme starts with 0 or the minimum number of joinpoints, which means a straight line fit on a log scale and tests whether more joinpoints must be added to the model (up to the pre-specified maximum number of join points) to better describe the trend in the data. The statistical significance of the change in trend is tested on the basis of a Monte Carlo permutation method (Kim et al, 2000). The number of joinpoint(s) are identified using the grid search method (Lerman, 1980) which allows a joinpoint to occur exactly at the year *t*. A grid is created for all possible positions of the joinpoint(s) or combination of joinpoint(s), the model is fitted for each possible position and that position is selected which minimises the sum of squared errors (SSE). In the present analysis, the minimum number of joinpoint(s) has been set to 0 while the maximum number of joinpoint(s) is set to 4.

Joinpoint regression analysis has frequently been used for analysing trend in mortality and morbidity from specific causes (Tyczynski and Berkel, 2005; Doucet, Rochette and Hamel, 2016; John and Hanke, 2015; Akinyede and Soyemi, 2016; Mogos et al, 2016; Chatenoud et al, 2015; Missikpode et al, 2015; Rea et al, 2017; Qiu et al, 2008; Puzo, Qin and Mehlum, 2016). It has also been used for estimating population parameters under changing population structure (Gillis and Edwards, 2019). Chaurasia (2020) has used joinpoint regression analysis to analyse the long-term trend in infant mortality in India. It has also been applied to analyse the trend in marital fertility (Chaurasia, 2020a), in understanding the rapid increase in life expectancy in Shanghai, China (Chen et al, 2018) and in analysing patterns and changes in life expectancy in China during 1990-2016 (Chen et al, 2020).

The second part of the analysis measures the contribution of the change in age-specific survival probabilities to the change in e_0_. Let the radix of the life table is *l*_0_. Suppose that the entire radix survives up to *N* years of age, then the total number of person years lived up to age *N* will be *N*\**l*_0_. If there is no death in the first year of life, the survival probability in the first year of life, *p*_0_, is 1 and total number of person years lived in the first year of life will be _1_*L*_0_=*l*_0_. If *p*_0_<1, then _1_*L*_0_ < *l*_0_ and person years lost in the first year of life is given by

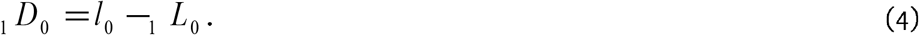

The persons years lost up to *N* years of age as the result of the mortality in the first year of life, therefore, is given by

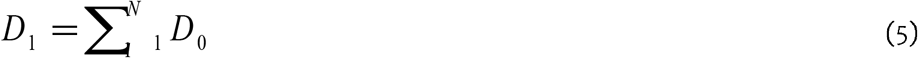

Similarly, the person years lost in the second year of life as the result of the probability of death in the second year of life is given by

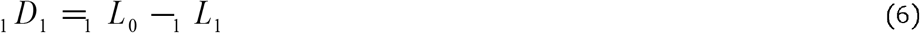

and the number of person years lost through all ages as the result of the mortality in the second year of life is given by

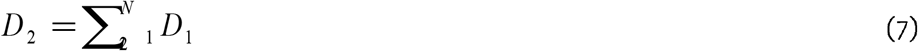

Finally, the total number of person years lost in the last year of the life table is given by

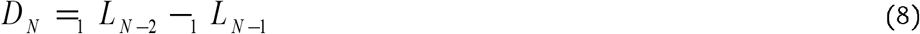

In other words, the total number of person-years lost due to the probability of death in different ages is given by

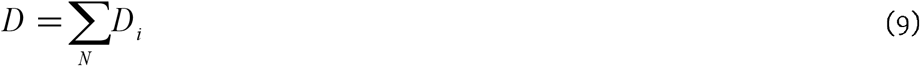

The expectation of life at birth, *e*_0_ may now be computed as

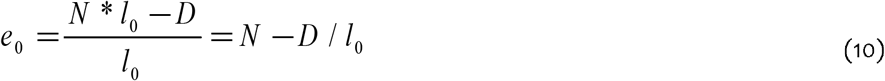

Equation (10) suggests that the change in *e*_0_ between two points in time, may now be decomposed as

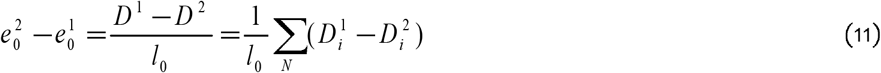

Equation (11) may also be used to decompose the change in the female-male gap or the gender gap and urban-rural gap or the residence gap in *e*_0_ over time. Equation (11) may also be used to analyse the relative contribution of age-specific survival experience in deciding the difference in *e*_0_ across different administrative units of sub-groups of the population.

## 4. Temporal Patterns in *e*_0_

The joinpoint regression analysis suggests that the trend in *e*_0_ in India changed three times between 2000 (1998-2000) and 2015 (2013-2017) (Table 1). The APC decreased considerably during 2002-2009 relative to 2000-2002; increased during 2009-2012 but again decreased during 2012-2015. As a result, *e*_0_ increased, on average, by 0.35 years per year during 2000-2002; by 0.32 years per year during 2002-2009; by 0.36 years per year during 2009-2012; and by only 0.28 years per year during 2012-2015. If the APC observed during 2000-2002 would have been sustained after 2002, *e*_0_ in India would have increased to more than 71 years by 2015. The deceleration in the increase in *e*_0_ during 2002-2009 and again during 2012-2015, appears to have resulted in a loss of more than two years in the increase e_0_ in the country during 2000-2015.

**Table 1:** Trend in the expectation of life at birth in India 2000 (1998-2002) through 2015 (2013-17). Net increase; average annual per cent change (AAPC); and Annual percent change (APC) in different time segments.

| Population groups | Net increase (years) | AAPC | APC in different time segments |  |  |  |  |  |  |  |  |  |  |  |  |  |  |  |
| --- | --- | --- | --- | --- | --- | --- | --- | --- | --- | --- | --- | --- | --- | --- | --- | --- | --- | --- |
|  |  |  | 2000 | 2001 | 2002 | 2003 | 2004 | 2005 | 2006 | 2007 | 2008 | 2009 | 2010 | 2011 | 2012 | 2013 | 2014 | 2015 |
| Person | 6.1 | 0.622* | 0.823* |  |  |  |  | 0.558* |  |  |  |  | 0.738* |  |  |  | 0.534* |  |
| Male | 5.9 | 0.613* | 0.670* |  |  |  |  | 0.448* |  |  |  |  |  |  | 0.750* |  |  |  |
| Female | 6.4 | 0.643* | 0.962* |  |  |  |  | 0.618* |  |  |  | 0.800* |  |  |  |  | 0.606* |  |
| Rural | 6.1 | 0.631* | 0.884* |  |  |  |  | 0.573* |  |  |  | 0.696* |  |  |  |  | 0.491* |  |
| Rural male | 6.6 | 0.601* | 0.653* |  |  |  |  | 0.460* |  |  |  |  |  |  | 0.683* |  |  |  |
| Rural female | 4.8 | 0.662* | 1.002* |  |  |  |  | 0.717* |  |  |  |  |  |  |  |  | 0.256* |  |
| Urban | 5.7 | 0.455* | 0.464* |  |  |  |  | 0.099 |  |  |  | 0.705* |  |  |  |  | 0.385* |  |
| Urban male | 5.1 | 0.498* | 0.592* |  |  |  |  | 0.100 |  |  |  | 0.715* |  |  |  |  | 0.506* |  |
| Urban female | 4.5 | 0.416* | 0.480* |  |  |  |  | 0.143 |  |  |  | 0.716* |  |  |  |  | 0.256* |  |

Source: Author's calculations
Remarks: The shaded cell indicates the joinpoint.
\* indicates that APC and AAPC are statistically significantly different from zero.

The joinpoint regression analysis also suggests that the trend in male *e*_0_ has been different from that in female *e*_0_. The trend in male *e*_0_ changed two times but the trend in female *e*_0_ changed three times during 2000-2015. The increase in male *e*_0_ accelerated during 2009-2015 so that male e_0_ increased at a rate of almost 0.49 years per year during this period. However, the increase in female *e*_0_ decelerated considerably during 2011-2015 so that female e_0_ increased at a rate of only 0.25 years per year during this period. During 2000-2011, female *e*_0_ in the country increased at an annual average rate of 0.730 per cent per year but male *e*_0_ increased at a rate of 0.563 per cent per year only. After 2011, however, male *e*_0_ increased at an average annual rate of 0.750 per cent per year whereas the average annual increase in female *e*_0_ decelerated to 0.606 per cent per year. As the result, the female-male gap in *e*_0_, which widened to 3.5 years by 2011, narrowed down considerably to 2.6 years by the year 2015.

The increase in *e*_0_ has also been comparatively faster in rural than in urban areas of the country. The trend in both rural and urban *e*_0_ changed three times but the years of change or joinpoints have been different. The rural *e*_0_ increased more rapidly than urban *e*_0_ during the period under reference except for the period 2007-2011. More importantly, increase in urban e_0_ stagnated during the period 2004-2007 as the APC, during this period, has not been found to be statistically significantly different from zero. As the result, the trend in urban-rural gap or the residence gap in *e*_0_ has been quite volatile. The residence gap in e_0_ decreased to 4.7 years in 2007; remained unchanged in 2008; increased to 4.9 years by the year 2011 but again decreased to 4.7 years by the year 2015.

The joinpoint regression analysis also suggests that the increase in *e*_0_ has been the fastest in rural females but the slowest in urban females during the period under reference. The rapid increase in rural female *e*_0_ has largely been due to the very rapid increase during 2000-2002 when it increased by more than 1 per cent per year, the most rapid in all population groups and in all time segments. However, after 2002, there has been considerable deceleration in the increase in rural female e_0_ and, during 2012-2015, it increased at annual average rate of only 0.253 per cent per year which was slower than the increase in rural male *e*_0_. In the urban areas, on the other hand, the increase in *e*_0_ stagnated in both males and female during 2004-2007. During 2012-2015, the rate of increase in *e*_0_ of rural females was almost the same as the rate of increase in e_0_ of urban females. On the other hand, the increase in rural male e_0_ accelerated but the increase in urban male e_0_ decelerated during this period so that the urban-rural gap in male *e*_0_ decreased during this period.

## 5. Decomposition of the Increase in *e*_0_

The *e*_0_ in India increased by around 6.1 years during the 15 years between 1998-2002 and 2013-2017. The increase in the person-years lived in the first year of life accounted for an increase of around 1.72 years while increase in person-years lived in 1-5 years of life accounted for an increase of 1.86 years so that increase in person-years lived in the first five years of life accounted for an increase of 3.58 years or more than 58 per cent of the increase in e_0_ (Table 2). Increase in person-years lived during 15-60 years of life accounted for an increase of 1.8 years or 30 per cent increase in *e*_0_. Increase in person-years lived in 60-75 years accounted for an increase of around 0.62 years or 10 per cent increase in *e*_0_ but the decrease in the person-years lived in the age group 75 years and above resulted in a decrease of around 0.85 years or 14 per cent decrease in *e*_0_. The average annual gain in e_0_ was the highest during 2009-2012 but the lowest during 2012-2015 because person-years lived in the age group 50-65 years decreased during 2012-2015 compared to 2009-2012. Another reason behind low average annual gain in *e*_0_ during 2012-2015 has been very slow improvement in the survival probability in the first five years of life leading to only a marginal increase in the person-years lived in this age group.

**Table 2:** Contribution of different age groups (years) to increase in e_0_ in India between 2000(1998-2000) and 2015 (2013-2017).

| Age | Combined population |  |  | Rural population |  |  | Urban population |  |  |
| --- | --- | --- | --- | --- | --- | --- | --- | --- | --- |
|  | Person | Male | Female | Person | Male | Female | Person | Male | Female |
| 0-1 | 1.72 | 1.77 | 1.61 | 1.72 | 1.75 | 1.63 | 1.22 | 1.38 | 1.01 |
| 1-5 | 1.86 | 1.70 | 2.10 | 2.08 | 1.90 | 2.35 | 0.89 | 0.83 | 0.99 |
| 5-10 | 0.70 | 0.58 | 0.82 | 0.76 | 0.63 | 0.89 | 0.41 | 0.34 | 0.47 |
| 10-15 | 0.30 | 0.26 | 0.33 | 0.33 | 0.29 | 0.37 | 0.15 | 0.13 | 0.17 |
| 15-20 | 0.21 | 0.18 | 0.25 | 0.24 | 0.20 | 0.29 | 0.10 | 0.09 | 0.12 |
| 20-25 | 0.29 | 0.21 | 0.37 | 0.33 | 0.24 | 0.44 | 0.14 | 0.11 | 0.18 |
| 25-30 | 0.31 | 0.24 | 0.38 | 0.33 | 0.26 | 0.41 | 0.22 | 0.17 | 0.27 |
| 30-35 | 0.26 | 0.24 | 0.29 | 0.25 | 0.22 | 0.29 | 0.24 | 0.26 | 0.21 |
| 35-40 | 0.21 | 0.20 | 0.22 | 0.20 | 0.18 | 0.22 | 0.19 | 0.23 | 0.14 |
| 40-45 | 0.15 | 0.17 | 0.14 | 0.12 | 0.13 | 0.11 | 0.18 | 0.20 | 0.15 |
| 45-50 | 0.13 | 0.13 | 0.13 | 0.09 | 0.09 | 0.09 | 0.18 | 0.20 | 0.15 |
| 50-55 | 0.11 | 0.18 | 0.04 | 0.03 | 0.12 | -0.06 | 0.24 | 0.27 | 0.18 |
| 55-60 | 0.13 | 0.19 | 0.05 | 0.01 | 0.07 | -0.07 | 0.34 | 0.38 | 0.26 |
| 60-65 | 0.20 | 0.24 | 0.17 | 0.10 | 0.10 | 0.10 | 0.39 | 0.47 | 0.30 |
| 65-70 | 0.29 | 0.38 | 0.21 | 0.21 | 0.30 | 0.15 | 0.45 | 0.56 | 0.36 |
| 70-75 | 0.13 | 0.11 | 0.15 | 0.10 | 0.08 | 0.13 | 0.20 | 0.19 | 0.22 |
| 75-80 | -0.10 | -0.16 | -0.03 | -0.09 | -0.15 | -0.03 | -0.09 | -0.19 | 0.02 |
| 80-85 | -0.35 | -0.38 | -0.33 | -0.36 | -0.37 | -0.34 | -0.29 | -0.31 | -0.26 |
| 85+ | -0.40 | -0.36 | -0.44 | -0.42 | -0.38 | -0.46 | -0.29 | -0.21 | -0.40 |
| Increase in $e_0$ | 6.15 | 5.89 | 6.44 | 6.05 | 5.65 | 6.50 | 4.85 | 5.10 | 4.53 |

Source: Author's calculations.

The relative contribution of the change in age-specific survival probabilities to the change in *e*_0_ has been different in females as compared to males. Almost 80 per cent of the increase in the female *e*_0_ during the period under reference is attributed to the increase in the number of person-years lived in the first 15 years of life. This proportion is only 70 per cent in males. On the other hand, increase in the number of person-years lived in the age group 60-75 years accounted for an increase of 0.73 years in male *e*_0_ but only 0.53 years increase in female *e*_0_. Finally, the decrease in the number of person-years lived in the age group 75 years and above accounted for a decrease of 0.81 years in female *e*_0_ but a decrease of 0.90 years in male *e*_0_. In 2000, female *e*_0_ was 2 years more than male *e*_0_ because females had survival advantage over males in the age groups 0-1 years 30-75 years and 85+ years. Males had survival advantage over females in the age group 1-30 years and in 75-85 years. In 2015, female *e*_0_ exceeded male *e*_0_ by almost 2.6 years because females had survival advantage over males in the age groups 15-80 years. However, males had survival advantage over females in the age groups 0-15 years and 80 years and above (Table 3).

**Table 3:** Decomposition of the gender gap (F-M) and residence gap (U-R) in e_0_ in India 2000 (1998-2000) and 2015 (2013-2017).

| Age | Gender gap (F-M) |  |  | Residence gap (U-R) |  |  |
| --- | --- | --- | --- | --- | --- | --- |
|  | 2000 | 2015 | Change | 2000 | 2015 | Change |
| 0-1 | 0.040 | -0.119 | -0.158 | 1.742 | 1.239 | -0.503 |
| 1-5 | -0.617 | -0.221 | 0.396 | 1.868 | 0.673 | -1.195 |
| 5-10 | -0.292 | -0.059 | 0.233 | 0.562 | 0.211 | -0.351 |
| 10-15 | -0.075 | -0.001 | 0.074 | 0.261 | 0.082 | -0.179 |
| 15-20 | -0.069 | 0.004 | 0.072 | 0.208 | 0.069 | -0.139 |
| 20-25 | -0.140 | 0.028 | 0.167 | 0.271 | 0.079 | -0.193 |
| 25-30 | -0.051 | 0.094 | 0.144 | 0.242 | 0.128 | -0.114 |
| 30-35 | 0.110 | 0.162 | 0.052 | 0.183 | 0.171 | -0.012 |
| 35-40 | 0.257 | 0.269 | 0.012 | 0.176 | 0.166 | -0.009 |
| 40-45 | 0.360 | 0.330 | -0.030 | 0.149 | 0.206 | 0.057 |
| 45-50 | 0.432 | 0.424 | -0.008 | 0.155 | 0.244 | 0.089 |
| 50-55 | 0.523 | 0.374 | -0.148 | 0.139 | 0.346 | 0.207 |
| 55-60 | 0.578 | 0.431 | -0.147 | 0.085 | 0.413 | 0.328 |
| 60-65 | 0.552 | 0.481 | -0.072 | 0.105 | 0.390 | 0.285 |
| 65-70 | 0.442 | 0.272 | -0.170 | 0.028 | 0.266 | 0.239 |
| 70-75 | 0.144 | 0.183 | 0.040 | 0.012 | 0.111 | 0.099 |
| 75-80 | -0.067 | 0.063 | 0.131 | -0.076 | -0.078 | -0.002 |
| 80-85 | -0.145 | -0.101 | 0.044 | -0.181 | -0.113 | 0.068 |
| 85+ | 0.066 | -0.018 | -0.083 | 0.027 | 0.154 | 0.127 |
| $e_0$ | 2.047 | 2.597 | 0.550 | 5.955 | 4.757 | -1.198 |

Source: Author's calculations.

On the other hand, increase in the person-years lived in the first five years of life accounted for almost two-third of the increase in *e*_0_ in the rural areas of the country but only around 43 per cent of the increase in *e*_0_ in the urban areas. Similarly, increase in the person-years lived in the age group 1-5 years of life accounted for more than 34 per cent of the increase in rural *e*_0_ but only around 18 per cent of the increase in urban *e*_0_. The urban e_0_ was higher than the rural e_0_ in 2000 as well as in 2015 because the urban population had survival advantage over rural population in the age group 0-75 years and in population aged 85 years and above. However, rural population had survival advantage over urban population in the age group 75-85 years in 2000 as well as in 2015 (Table 3).

The relative contribution of the change in survival experience in different age groups to the change in e_0_ has been different in different time segments of the period 2000-2015 identified through the joinpoint regression analysis. The deceleration in the in crease in male e_0_ during 2004-2009 was primarily due to the decrease in person-years lived in the age group 40-55 years and 65 years and above. During this period, the contribution of the increase in the person-years lived in the first 15 years of life was higher than the increase in e_0_ during this period (Table 4). On the other hand, the deceleration in the decrease in female e_0_ during 2011-2015 was primarily due to the decrease in the person-years lived in the age group 40-65 years and 85 years and above.

**Table 4:**
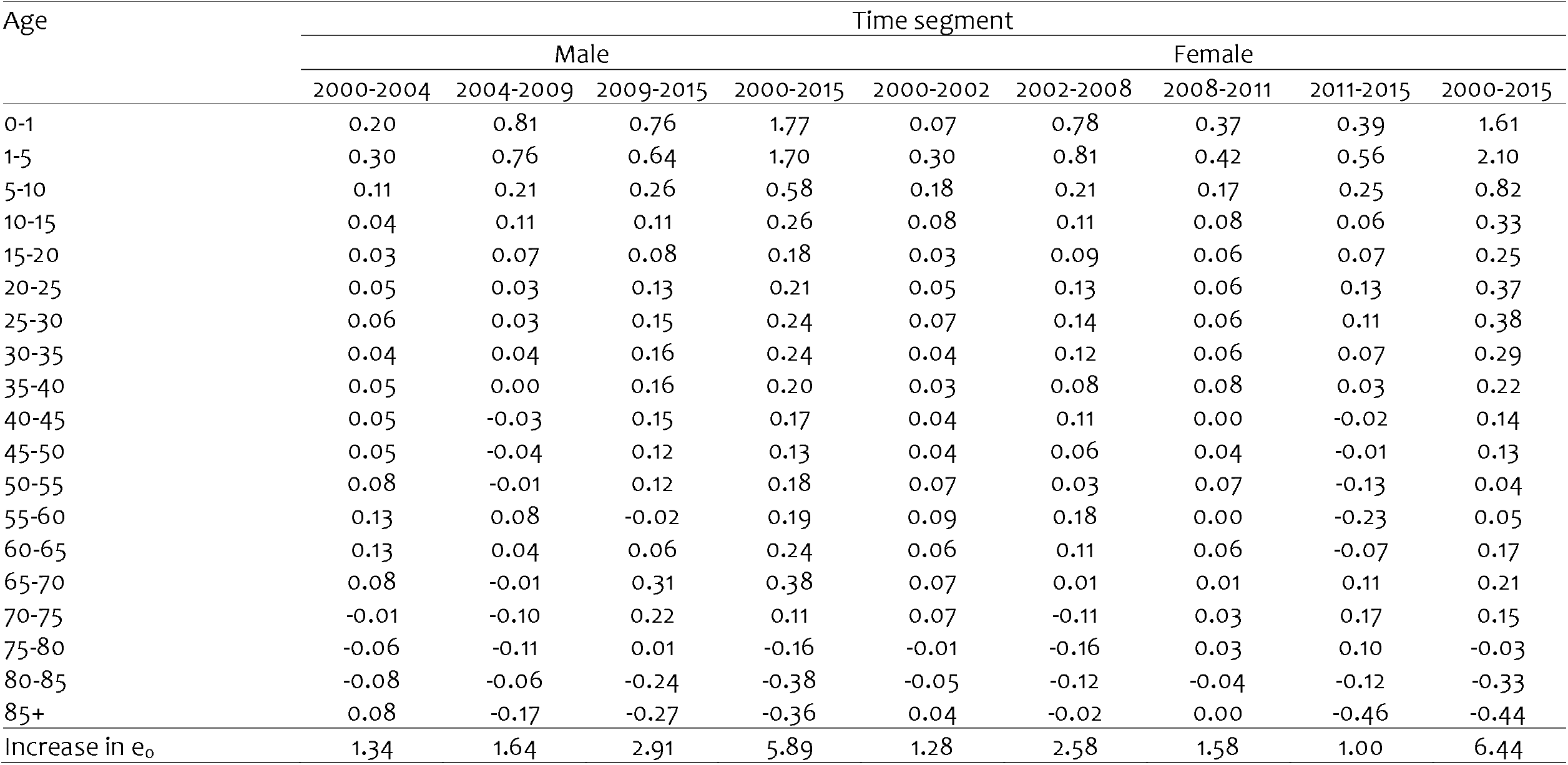
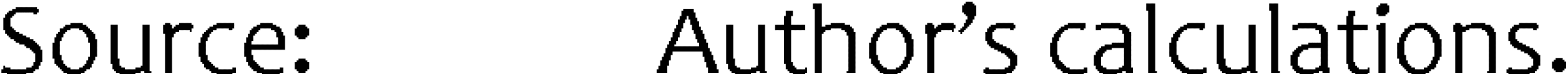
Decomposition of the increase in male and female e_0_ in India in different time segments identified through joinpoint regression analysis.

| Age | Time segment |  |  |  |  |  |  |  |  |
| --- | --- | --- | --- | --- | --- | --- | --- | --- | --- |
|  | Male |  |  |  | Female |  |  |  |  |
|  | 2000-2004 | 2004-2009 | 2009-2015 | 2000-2015 | 2000-2002 | 2002-2008 | 2008-2011 | 2011-2015 | 2000-2015 |
| 0-1 | 0.20 | 0.81 | 0.76 | 1.77 | 0.07 | 0.78 | 0.37 | 0.39 | 1.61 |
| 1-5 | 0.30 | 0.76 | 0.64 | 1.70 | 0.30 | 0.81 | 0.42 | 0.56 | 2.10 |
| 5-10 | 0.11 | 0.21 | 0.26 | 0.58 | 0.18 | 0.21 | 0.17 | 0.25 | 0.82 |
| 10-15 | 0.04 | 0.11 | 0.11 | 0.26 | 0.08 | 0.11 | 0.08 | 0.06 | 0.33 |
| 15-20 | 0.03 | 0.07 | 0.08 | 0.18 | 0.03 | 0.09 | 0.06 | 0.07 | 0.25 |
| 20-25 | 0.05 | 0.03 | 0.13 | 0.21 | 0.05 | 0.13 | 0.06 | 0.13 | 0.37 |
| 25-30 | 0.06 | 0.03 | 0.15 | 0.24 | 0.07 | 0.14 | 0.06 | 0.11 | 0.38 |
| 30-35 | 0.04 | 0.04 | 0.16 | 0.24 | 0.04 | 0.12 | 0.06 | 0.07 | 0.29 |
| 35-40 | 0.05 | 0.00 | 0.16 | 0.20 | 0.03 | 0.08 | 0.08 | 0.03 | 0.22 |
| 40-45 | 0.05 | -0.03 | 0.15 | 0.17 | 0.04 | 0.11 | 0.00 | -0.02 | 0.14 |
| 45-50 | 0.05 | -0.04 | 0.12 | 0.13 | 0.04 | 0.06 | 0.04 | -0.01 | 0.13 |
| 50-55 | 0.08 | -0.01 | 0.12 | 0.18 | 0.07 | 0.03 | 0.07 | -0.13 | 0.04 |
| 55-60 | 0.13 | 0.08 | -0.02 | 0.19 | 0.09 | 0.18 | 0.00 | -0.23 | 0.05 |
| 60-65 | 0.13 | 0.04 | 0.06 | 0.24 | 0.06 | 0.11 | 0.06 | -0.07 | 0.17 |
| 65-70 | 0.08 | -0.01 | 0.31 | 0.38 | 0.07 | 0.01 | 0.01 | 0.11 | 0.21 |
| 70-75 | -0.01 | -0.10 | 0.22 | 0.11 | 0.07 | -0.11 | 0.03 | 0.17 | 0.15 |
| 75-80 | -0.06 | -0.11 | 0.01 | -0.16 | -0.01 | -0.16 | 0.03 | 0.10 | -0.03 |
| 80-85 | -0.08 | -0.06 | -0.24 | -0.38 | -0.05 | -0.12 | -0.04 | -0.12 | -0.33 |
| 85+ | 0.08 | -0.17 | -0.27 | -0.36 | 0.04 | -0.02 | 0.00 | -0.46 | -0.44 |
| Increase in $e_0$ | 1.34 | 1.64 | 2.91 | 5.89 | 1.28 | 2.58 | 1.58 | 1.00 | 6.44 |

Source: Author's calculations.

## 6. Inter-state Variations

The trend in *e*_0_ has varied across states for which official life tables are available for the year 2000 (1998-2002) and 2015 (2013-2017) (Table 5). In Andhra Pradesh and Kerala, the trend in *e*_0_ changed four times whereas Madhya Pradesh is the only state where there has been no change in the trend or *e*_0_ increased linearly (on a log scale) during the period 2000-2015. In majority of the states, however, the trend in *e*_0_ changed three times. The increase in *e*_0_ has been the fastest in Odisha but the slowest in Kerala. Odisha is the only state where female *e*_0_ increased by more than 9 years during 2000-2015 whereas Kerala is the only state where *e*_0_ increased by less than 4 years. Inter-state variance in *e*_0_, however, decreased over time indicating sigma-convergence in *e*_0_. There are seven states where APC was not statistically significantly different from zero for sometimes during 2000-2015 reflecting that the increase in e_0_ stagnated in this time-segment. In Andhra Pradesh, the increase in *e*_0_ stagnated in three out of five time-segments. In Kerala and Punjab, increase in *e*_0_ stagnated in two time-segments while *e*_0_ stagnated in one time segment in Bihar, Haryana, Jammu and Kashmir and Maharashtra. The annual per cent increase in *e*_0_ (APC) was the fastest in Jammu and Kashmir during 2000-2004 but the slowest in Kerala during 2013-2015. In most of the states, increase in *e*_0_ decelerated during the later years of the period 2000-15 as compared to the earlier years of the period with the exception of only two states - Assam and Odisha. The deceleration in the increase in *e*_0_ has been particularly marked in female *e*_0_. Odisha is the only state where increase in female *e*_0_ did not decelerate during the period under reference whereas, in 9 states, increase in female *e*_0_ stagnated at least during 2013-2015 as the APC was not statistically significantly different from 0 during this period. By contrast, there is no state where the increase in male *e*_0_ stagnated in recent years. In all states, the volatility in the trend has also been found to be less in male *e*_0_ compared to female *e*_0_.

**Table 5:**
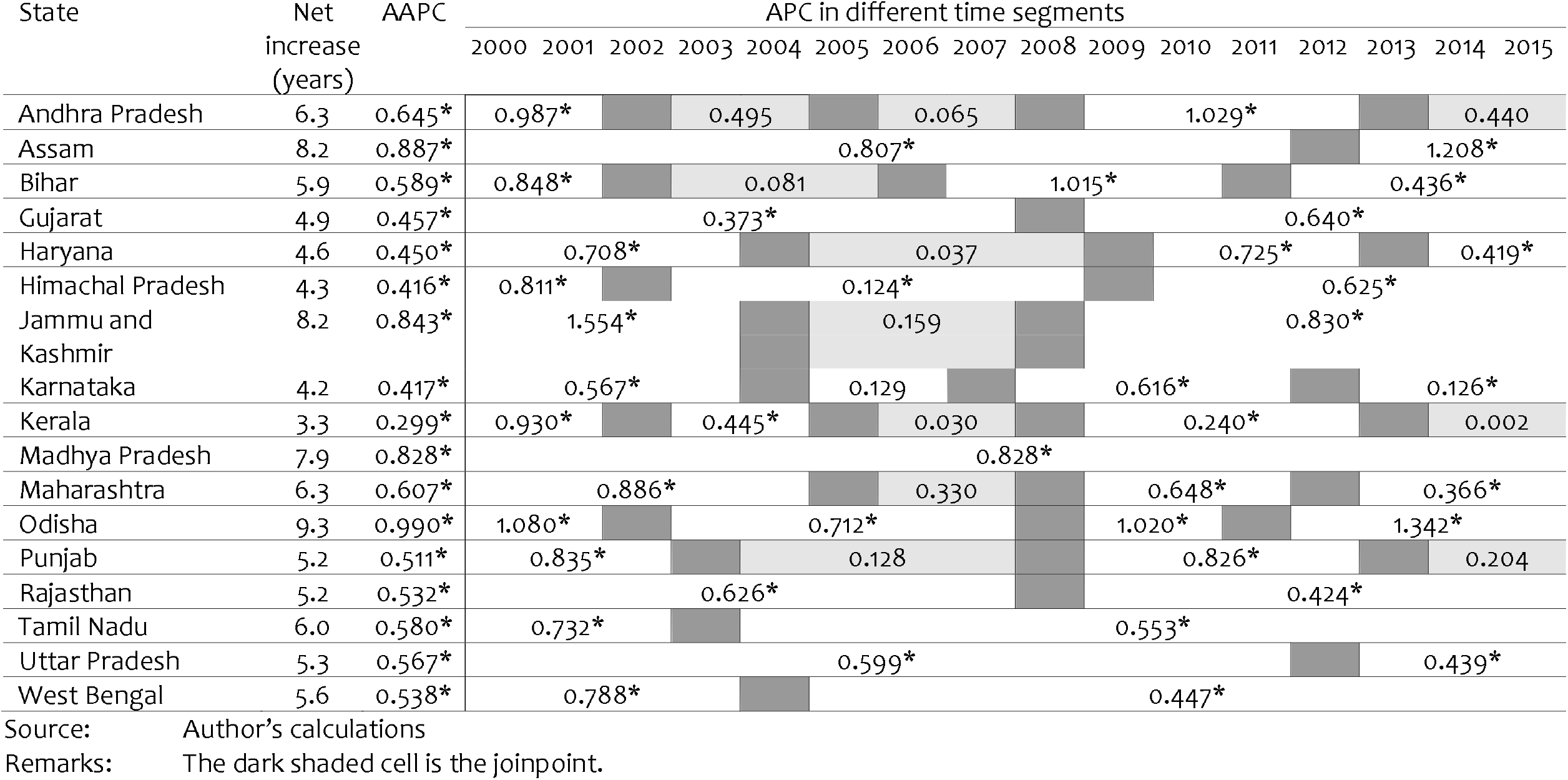
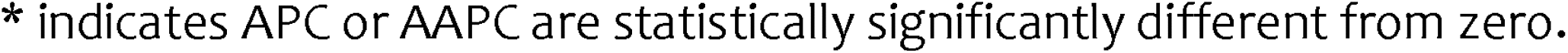
Trend in e_0_ in selected states, 1998-2002 (circa 2000) through 2013-17 (circa 2015). Net increase; average annual per cent change (AAPC); and Annual percent change (APC) in different time segments.

| State | Net<br>increase<br>(years) | AAPC | APC in different time segments |  |  |  |  |  |  |  |  |  |  |  |  |  |  |
| --- | --- | --- | --- | --- | --- | --- | --- | --- | --- | --- | --- | --- | --- | --- | --- | --- | --- |
|  |  |  | 2000 | 2001 | 2002 | 2003 | 2004 | 2005 | 2006 | 2007 | 2008 | 2009 | 2010 | 2011 | 2012 | 2013 | 2014 |
| Andhra Pradesh | 6.3 | 0.645* | 0.987* |  |  | 0.495 |  | 0.065 |  |  | 1.029* |  |  |  |  |  | 0.440 |
| Assam | 8.2 | 0.887* | 0.807* |  |  |  |  |  |  |  |  |  |  |  | 1.208* |  |  |
| Bihar | 5.9 | 0.589* | 0.848* |  |  | 0.081 |  |  |  | 1.015* |  |  |  |  | 0.436* |  |  |
| Gujarat | 4.9 | 0.457* | 0.373* |  |  |  |  |  |  |  |  | 0.640* |  |  |  |  |  |
| Haryana | 4.6 | 0.450* | 0.708* |  |  |  |  | 0.037 |  |  |  |  | 0.725* |  |  |  | 0.419* |
| Himachal Pradesh | 4.3 | 0.416* | 0.811* |  |  | 0.124* |  |  |  |  |  |  | 0.625* |  |  |  |  |
| Jammu and<br>Kashmir | 8.2 | 0.843* | 1.554* |  |  |  |  | 0.159 |  |  |  |  | 0.830* |  |  |  |  |
| Karnataka | 4.2 | 0.417* | 0.567* |  |  |  |  | 0.129 |  |  | 0.616* |  |  |  |  |  | 0.126* |
| Kerala | 3.3 | 0.299* | 0.930* |  |  | 0.445* |  | 0.030 |  |  | 0.240* |  |  |  |  |  | 0.002 |
| Madhya Pradesh | 7.9 | 0.828* | 0.828* |  |  |  |  |  |  |  |  |  |  |  |  |  |  |
| Maharashtra | 6.3 | 0.607* | 0.886* |  |  |  |  | 0.330 |  |  | 0.648* |  |  |  |  | 0.366* |  |
| Odisha | 9.3 | 0.990* | 1.080* |  |  | 0.712* |  |  |  |  |  | 1.020* |  |  | 1.342* |  |  |
| Punjab | 5.2 | 0.511* | 0.835* |  |  |  | 0.128 |  |  |  |  | 0.826* |  |  |  |  | 0.204 |
| Rajasthan | 5.2 | 0.532* | 0.626* |  |  |  |  |  |  |  | 0.424* |  |  |  |  |  |  |
| Tamil Nadu | 6.0 | 0.580* | 0.732* |  |  |  | 0.553* |  |  |  |  |  |  |  |  |  |  |
| Uttar Pradesh | 5.3 | 0.567* | 0.599* |  |  |  |  |  |  |  |  |  |  |  | 0.439* |  |  |
| West Bengal | 5.6 | 0.538* | 0.788* |  |  |  |  | 0.447* |  |  |  |  |  |  |  |  |  |

Source: Author's calculations
Remarks: The dark shaded cell is the joinpoint.
\* indicates APC or AAPC are statistically significantly different from zero.

**Table 5:**
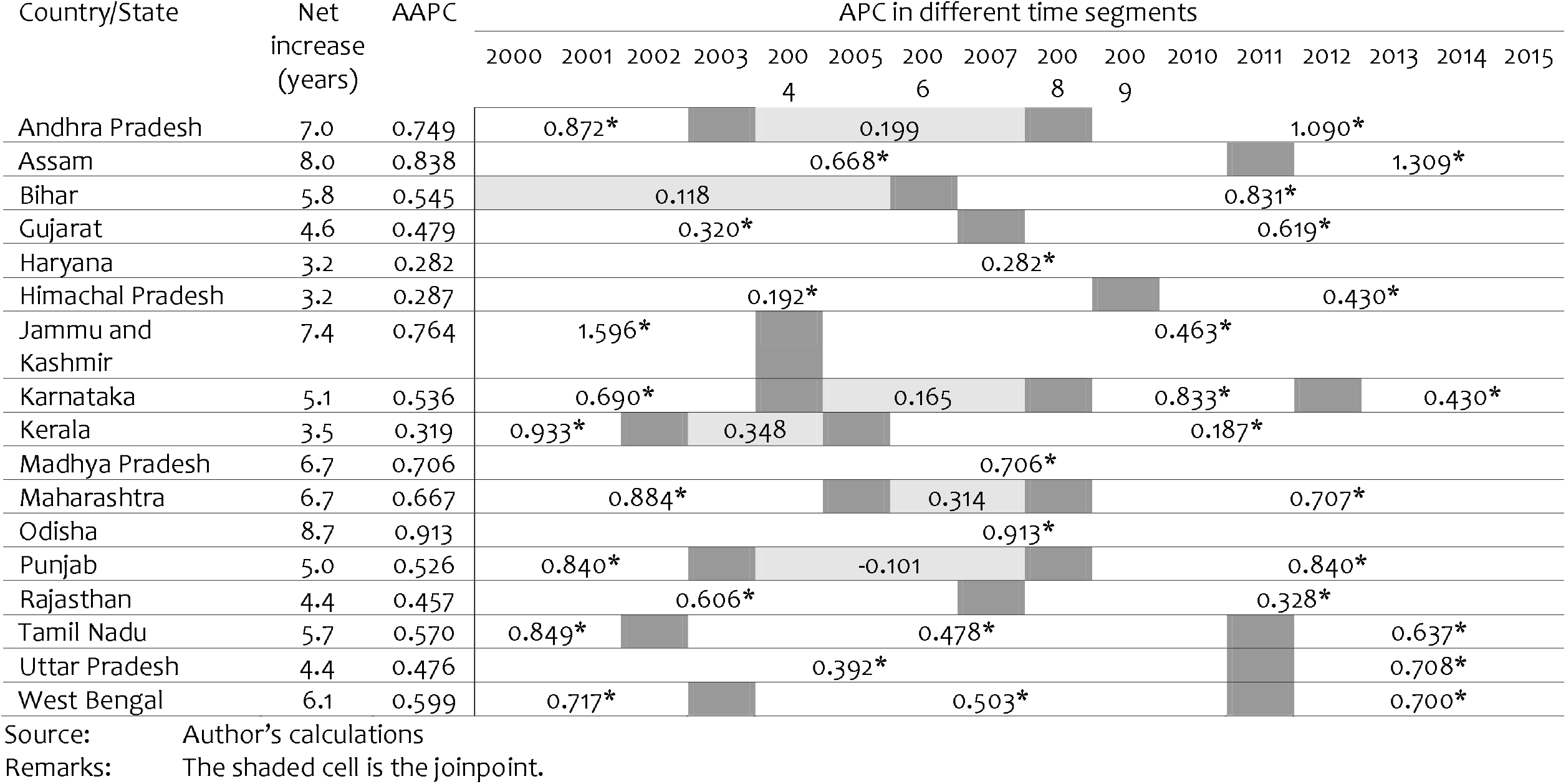
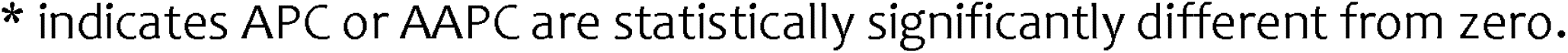
Trend in male e_0_ in selected states, 2000(1998-2002) through 2015 (2013-17). Net increase; average annual per cent change (AAPC); and Annual percent change (APC) in different time segments.

| Country/State | Net<br>increase<br>(years) | AAPC | APC in different time segments |  |  |  |  |  |  |  |  |  |  |  |  |  |  |  |
| --- | --- | --- | --- | --- | --- | --- | --- | --- | --- | --- | --- | --- | --- | --- | --- | --- | --- | --- |
|  |  |  | 2000 | 2001 | 2002 | 2003 | 2004 | 2005 | 2006 | 2007 | 2008 | 2009 | 2010 | 2011 | 2012 | 2013 | 2014 | 2015 |
| Andhra Pradesh | 7.0 | 0.749 | 0.872* |  |  |  | 0.199 |  |  |  |  |  | 1.090* |  |  |  |  |  |
| Assam | 8.0 | 0.838 | 0.668* |  |  |  |  |  |  |  |  |  |  |  | 1.309* |  |  |  |
| Bihar | 5.8 | 0.545 | 0.118 |  |  |  |  |  |  |  |  |  | 0.831* |  |  |  |  |  |
| Gujarat | 4.6 | 0.479 | 0.320* |  |  |  |  |  |  |  | 0.619* |  |  |  |  |  |  |  |
| Haryana | 3.2 | 0.282 |  |  |  |  |  |  | 0.282* |  |  |  |  |  |  |  |  |  |
| Himachal Pradesh | 3.2 | 0.287 |  |  |  |  | 0.192* |  |  |  |  |  |  |  | 0.430* |  |  |  |
| Jammu and<br>Kashmir | 7.4 | 0.764 | 1.596* |  |  |  |  |  | 0.463* |  |  |  |  |  |  |  |  |  |
| Karnataka | 5.1 | 0.536 | 0.690* |  |  |  |  |  | 0.165 |  |  |  | 0.833* |  |  |  | 0.430* |  |
| Kerala | 3.5 | 0.319 | 0.933* |  |  |  | 0.348 |  |  |  | 0.187* |  |  |  |  |  |  |  |
| Madhya Pradesh | 6.7 | 0.706 |  |  |  |  |  |  |  |  |  |  | 0.706* |  |  |  |  |  |
| Maharashtra | 6.7 | 0.667 | 0.884* |  |  |  |  |  | 0.314 |  |  |  | 0.707* |  |  |  |  |  |
| Odisha | 8.7 | 0.913 |  |  |  |  |  |  |  |  |  |  | 0.913* |  |  |  |  |  |
| Punjab | 5.0 | 0.526 | 0.840* |  |  |  | -0.101 |  |  |  |  |  | 0.840* |  |  |  |  |  |
| Rajasthan | 4.4 | 0.457 | 0.606* |  |  |  |  |  |  |  | 0.328* |  |  |  |  |  |  |  |
| Tamil Nadu | 5.7 | 0.570 | 0.849* |  |  |  | 0.478* |  |  |  |  |  |  |  | 0.637* |  |  |  |
| Uttar Pradesh | 4.4 | 0.476 | 0.392* |  |  |  |  |  |  |  |  |  |  |  | 0.708* |  |  |  |
| West Bengal | 6.1 | 0.599 | 0.717* |  |  |  | 0.503* |  |  |  |  |  |  |  | 0.700* |  |  |  |

Source: Author's calculations
Remarks: The shaded cell is the joinpoint.
\* indicates APC or AAPC are statistically significantly different from zero.

**Table 5:**
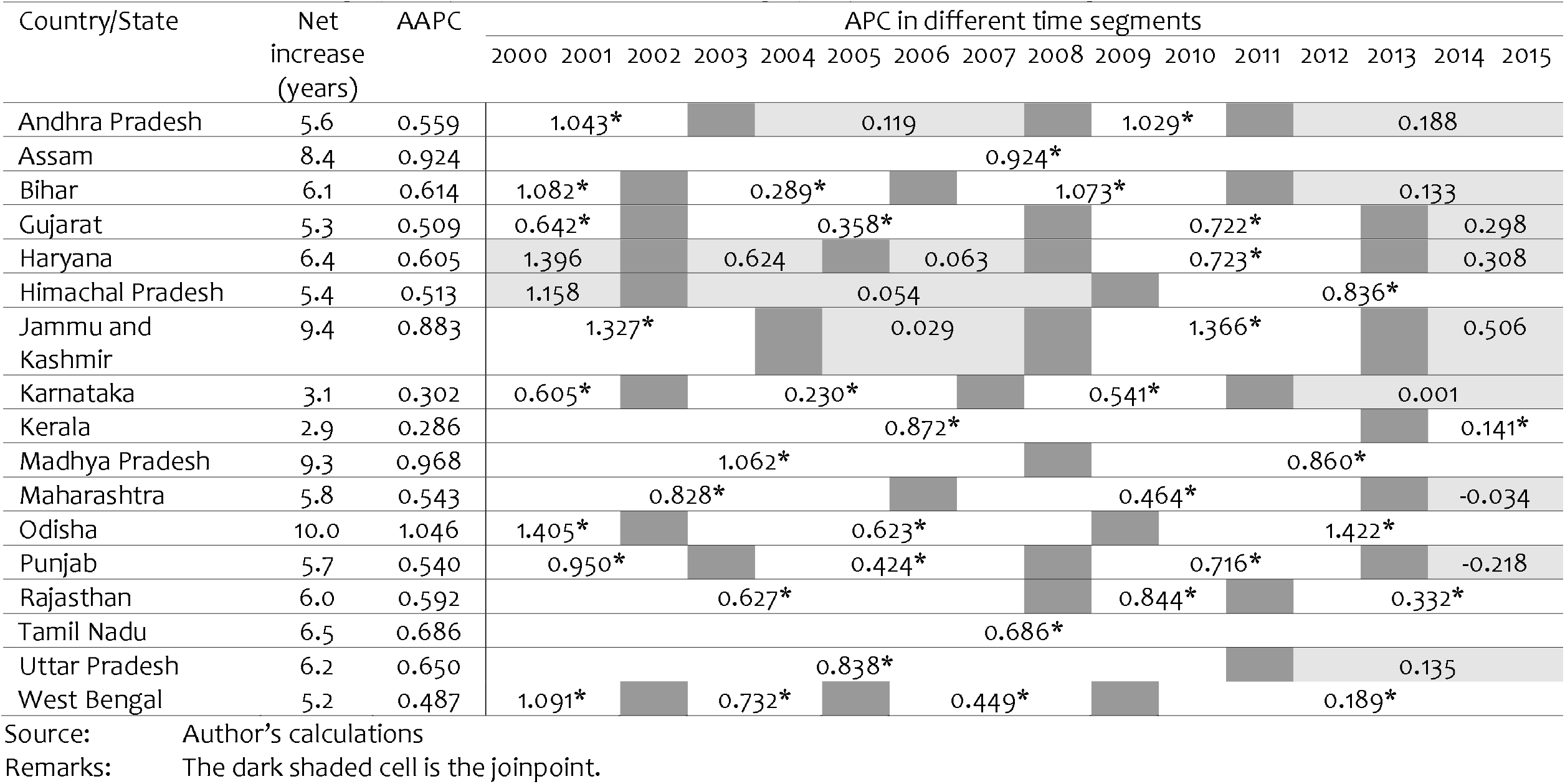
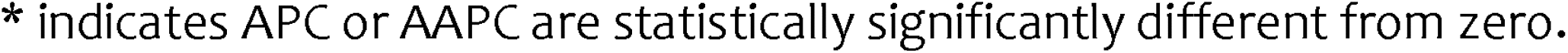
Trend in female e_0_ in selected states, 2000 (1998-2002) through 2015 (2013-17). Net increase; average annual per cent change (AAPC); and Annual percent change (APC) in different time segments.

| Country/State | Net increase (years) | AAPC | APC in different time segments |  |  |  |  |  |  |  |  |  |  |  |  |  |  |  |
| --- | --- | --- | --- | --- | --- | --- | --- | --- | --- | --- | --- | --- | --- | --- | --- | --- | --- | --- |
|  |  |  | 2000 | 2001 | 2002 | 2003 | 2004 | 2005 | 2006 | 2007 | 2008 | 2009 | 2010 | 2011 | 2012 | 2013 | 2014 | 2015 |
| Andhra Pradesh | 5.6 | 0.559 | 1.043* |  |  |  | 0.119 |  |  |  | 1.029* |  |  | 0.188 |  |  |  |  |
| Assam | 8.4 | 0.924 | 0.924* |  |  |  |  |  |  |  |  |  |  |  |  |  |  |  |
| Bihar | 6.1 | 0.614 | 1.082* |  |  | 0.289* |  |  |  | 1.073* |  |  |  | 0.133 |  |  |  |  |
| Gujarat | 5.3 | 0.509 | 0.642* |  |  | 0.358* |  |  |  |  | 0.722* |  |  |  |  |  | 0.298 |  |
| Haryana | 6.4 | 0.605 | 1.396 |  |  | 0.624 |  |  | 0.063 |  |  | 0.723* |  |  |  |  | 0.308 |  |
| Himachal Pradesh | 5.4 | 0.513 | 1.158 |  |  | 0.054 |  |  |  |  |  | 0.836* |  |  |  |  |  |  |
| Jammu and Kashmir | 9.4 | 0.883 | 1.327* |  |  |  | 0.029 |  |  |  | 1.366* |  |  |  |  |  | 0.506 |  |
| Karnataka | 3.1 | 0.302 | 0.605* |  |  | 0.230* |  |  |  |  | 0.541* |  |  |  | 0.001 |  |  |  |
| Kerala | 2.9 | 0.286 |  |  |  |  |  |  | 0.872* |  |  |  |  |  |  |  | 0.141* |  |
| Madhya Pradesh | 9.3 | 0.968 |  |  |  |  |  | 1.062* |  |  |  | 0.860* |  |  |  |  |  |  |
| Maharashtra | 5.8 | 0.543 |  |  |  |  | 0.828* |  |  |  | 0.464* |  |  |  |  |  | -0.034 |  |
| Odisha | 10.0 | 1.046 | 1.405* |  |  |  |  |  |  | 0.623* |  |  |  |  |  | 1.422* |  |  |
| Punjab | 5.7 | 0.540 | 0.950* |  |  |  |  |  |  |  | 0.424* |  |  | 0.716* |  |  |  | -0.218 |
| Rajasthan | 6.0 | 0.592 |  |  |  |  |  | 0.627* |  |  |  | 0.844* |  |  | 0.332* |  |  |  |
| Tamil Nadu | 6.5 | 0.686 | 0.686* |  |  |  |  |  |  |  |  |  |  |  |  |  |  |  |
| Uttar Pradesh | 6.2 | 0.650 |  |  |  |  |  | 0.838* |  |  |  |  |  |  |  | 0.135 |  |  |
| West Bengal | 5.2 | 0.487 | 1.091* |  |  | 0.732* |  |  | 0.449* |  |  |  |  |  | 0.189* |  |  |  |

Source: Author's calculations
Remarks: The dark shaded cell is the joinpoint.
\* indicates APC or AAPC are statistically significantly different from zero.

The relative contribution of the change in the survival experience in different age-groups to the change in *e*_0_ has also been different in different states (Table 6). In most of the states, increase in *e*_0_ has primarily been the result of the improvement in the survival experience in the first five years of life. Notable exceptions to this general pattern are Jammu and Kashmir and Kerala. Similarly, decrease in the number of person-years lived in the age group 75 years and above has accounted for a decrease, instead increase, in *e*_0_ in most of the states. There are only four states - Haryana, Jammu and Kashmir, Kerala and Punjab - where number of person-years lived in the age group 75 years and above increased during the period under reference and, therefore, the survival experience in this age group, has contributed to the increase in *e*_0_. In Haryana, the number of person-years lived in the age group 60-75 years decreased, instead increased, in 2015 compared to 2000. Similarly, the number of person years lived decreased, instead increased, in the age group 40-65 years in Rajasthan and in the age group 45-65 years in Utter Pradesh. The decrease in the person-years lived in these age groups has contributed towards slowing down the increase in *e*_0_ in these states during the period under reference.

**Table 6:**
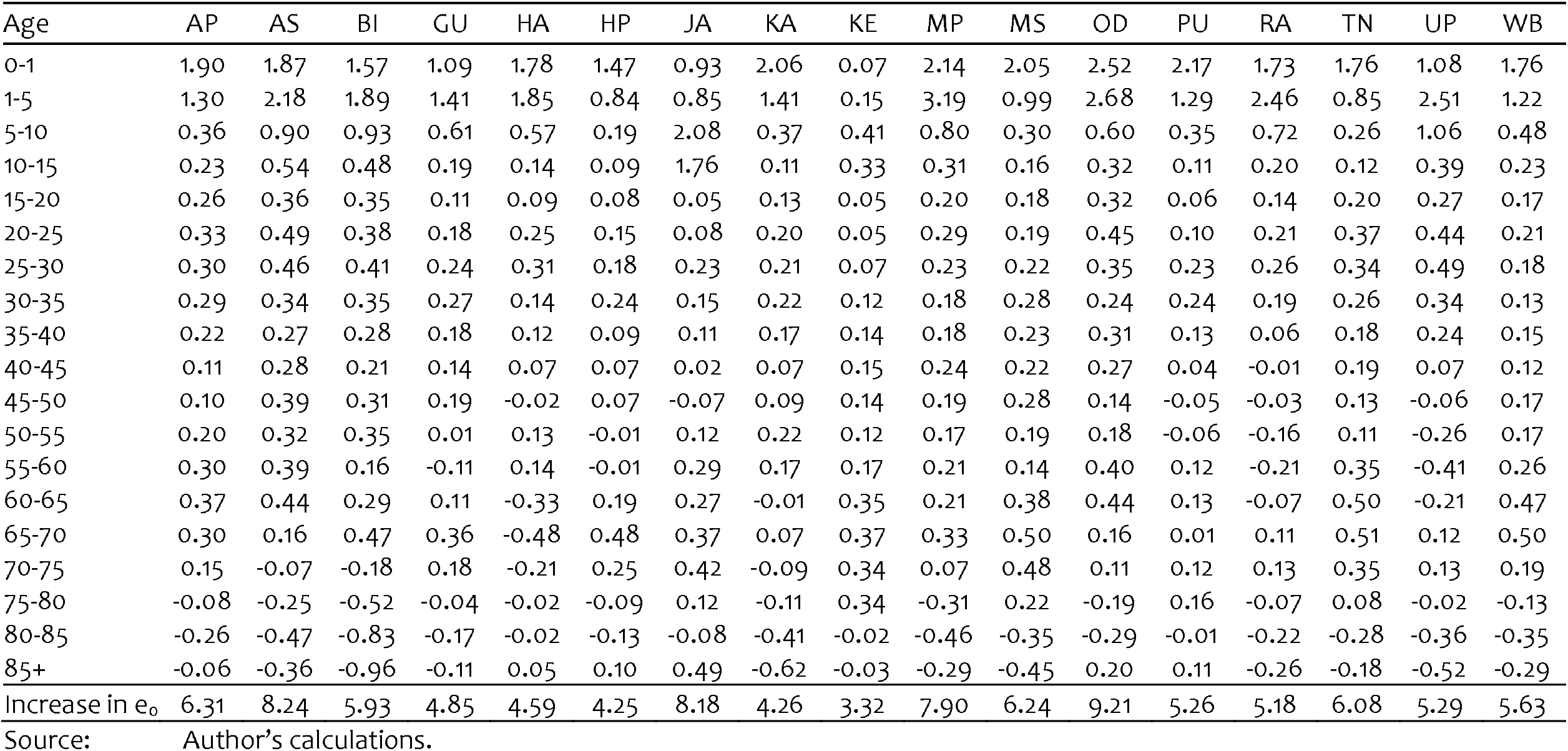
Contribution of different age groups (years) to increase in e_0_ in different states between 2000 and

## 7. Discussions and Conclusions

The present analysis reveals unsatisfactory trend in *e*_0_ in India, in its different population groups and in 17 states during the period 2000 (1998-2002) through 2015 (2013-2017). The trend in *e*_0_ is characterised by substantial degree of volatility. Most importantly, there has been considerable degree of deceleration in the increase in e_0_ which has been quite marked in females so that the increase in e_0_ slowed down considerably over time. The increase in *e*_0_ has also decelerated in most states of the country. The primary reason for the slowdown in the increase in *e*_0_ has been a rapid deceleration and even stagnation in the increase in female *e*_0_.

The analysis also reveals that the increase *e*_0_ stagnated in the urban areas of the country during 2003-2007 and this stagnation has primarily been responsible for relatively slower increase in urban *e*_0_ as compared to the increase in rural *e*_0_. Unlike the urban areas, there has been no stagnation in the increase in rural *e*_0_. The urban-rural gap in *e*_0_ narrowed down again in the recent past as the increase in urban *e*_0_ decelerated again. The deceleration in the increase in urban *e*_0_ has not been confined to a particular sex but is evident in both sexes.

The increase in *e*_0_ in the country during the period under reference has largely been the result of the improvement in the survival experience in the first fifteen years of life, although, this contribution has varied in different time segments and, in recent years, there has contribution has decreased substantially after 2002-2009. On the other hand, the number of person-years lived in the age group 75 years and above has also decreased despite the increase in the probability of survival in the age group 75-85 years. It appears that improvement in survival probability has not been large enough to ensure a decrease in the number of deaths and hence increase in the number of person-years lived in these age groups so that person-years lived in these age groups decreased.

Perhaps the most revealing observation of the present analysis is the considerable deceleration in the increase in female *e*_0_ in recent years. This deceleration appears to be the result of the decrease in the person-years lived in the age group 40-65 years despite an increase in the survival probability in the age group. Obviously, the increase in the female survival probability in this age group has not been large enough to ensure the decrease in the number of deaths and hence an increase in the number of person-years lived in this age group. In order to ensure that improvement in survival probability actually results in the increase in the person-years lived and hence contributes to the increase in *e*_0_, it is imperative that the improvement in survival probability is sufficiently large enough to ensure a decrease in the number of deaths and hence an increase in the number of person-years lived in the age group.

Reasons for the volatile trend in *e*_0_ and the considerable slowdown in the increase in *e*_0_, especially in females and in the urban areas of the country are not known at present. The period 2000-2015 has been a period of rapid, unprecedented economic growth in India. At the same time, there has been attempts to introduce a paradigm shift in the delivery of health care services so as to improve the efficiency of the health care delivery system in meeting the health and survival needs of the people of the country. It, however, appears that there has been little impact of rapid economic growth and paradigm shift in the health care delivery system on the improvement in population health as reflected in the trend in the expectation of life at birth. It appears that the dividends of the rapid economic growth that India has witnessed during this period could not be translated into social and human progress that is necessary for improving population health. It also appears that the paradigm shift in the delivery of health care services attempted first through National Rural Health Mission and subsequently through National Health Mission has contributed only marginally in improving the efficiency and effectiveness of health care delivery services in meeting the health care needs of the people. There has, however, been some degree of convergence in the expectation of life at birth across the states of the country.

The available evidence also confirms that the country has not been able to achieve the goal of an e_0_ of 75 years by the year 2015 that was set in the Plan of Action adopted at the 1994 International Conference on Population and Development at Cairo and to which India is a signatory. Interestingly, instead of acknowledging the inability to achieve this cherished goal and to identify and remedy the reasons for the sluggish improvement, the National Health Policy 2017 has significantly scaled down the goal to increasing the expectation of life at birth to 70 years by the year 2025. The current trend in the expectation of life at birth suggests the goal set in the National Health Policy 2017 will be achieved even without any acceleration in the current rate of increase in *e*_0_. If the expectation of life at birth is any indication, then, population in India remains amongst the poorest in the world and it will remain so in the coming next 10 years if one goes by the goal set in the National Health Policy 2017.

## Data Availability

All data available free online.

https://www.censusindia.gov.in

## References

Akinyede O, Soyemi K (2016) Joinpoint regression analysis of pertussis crude incidence rates, Illinois, 1990-2014. American Journal of Infection Control 44(12):1732–3.

Chaurasia AR (2010) Mortality transition in India: 1970-2005. Asian Population Studies 6(01): 47–68

Chaurasia AR (2020) Long-term trend in infant mortality in India: a joinpoint regression analysis for 1981-2018. Indian Journal of Human Development 14(3): 394–406.

Chaurasia AR (2020a) Fertility transition in currently married reproductive age women in India: 1985-2017. https://doi.org/10.1101/2020.07.16.20155176

Chatenoud L, Garavello W, Pagan E, Bertuccio P, Gallus S, La Vecchia C, Negri E, Bosetti C (2015) Laryngeal cancer mortality trends in European countries. International Journal of Cancer 842: 833–42.

Chen H, Qian Y, Dong Y, Yang Z, Guo L, Liu J, Shen Q, Wang L (2020) Patterns and changes in life expectancy in China, 1990-2016. PLoS ONE 15(4): e0231007. https://doi.org/10.1371/journal.pone.0231007

Chen H, Hao L, Yang C, Yan B, Sun Q, Sun L, Chen H, Chen (2018) Understanding the rapid increase in life expectancy in Shanghai, China: a population-based retrospective analysis. BMC Public Health 18:256. DOI 10.1186/s12889-018-5112-7

Clegg LX, Hankey BF, Tiwari R, Feuer EJ, Edwards BK (2009) Estimating average annual percent change in trend analysis. Statistics in Medicine 20 (29): 3670–82.

Doucet M, Rochette, Hamel D (2016) Prevalence and mortality trends in chronic obstructive pulmonary disease over 2001 to 2011: a public health point of view of the burden. Canadian Respiratory Journal 2016:1–10.

Gillis D, Edwards BPM (2019) The utility of joinpoint regression for estimating population parameters given changes in population structure. Heliyon 5: e02515.

Government of India (1971) Sample Registration of Births and Deaths in India 1969-70. New Delhi, Registrar General.

Government of India (1983) Report on intensive enquiry conducted in a subsample of SRS units (1980-81). Occasional Paper No. 2. New Delhi, Registrar General.

Government of India (1988) Report on intensive enquiry conducted in a sub-sample of SRS units. Occasional Paper 1 of 1988. New Delhi, Registrar General.

Government of India (2000) National Population Policy 2000. New Delhi, Ministry of Health and Family Welfare.

Government of India (2002) National Health Policy 2002. New Delhi, Ministry of Health and Family Welfare.

Government of India (2005) National Rural Health Mission. Framework of Implementation. New Delhi, Ministry of Health and Family Welfare.

Government of India (2013) National Health Mission. Framework of Implementation. New Delhi, Ministry of Health and Family Welfare.

Government of India (2017) National Health Policy 2017. New Delhi, Ministry of Health and Family Welfare.

Government of India (2018) Economic Survey 2017-18. New Delhi, Ministry of Finance. Department of Economic Affairs.

Government of India (2019) SRS Based Abridged Life Tables 2013-2017. New Delhi, Registrar General and Census Commissioner of India.

John U, Hanke M (2015) Liver cirrhosis mortality, alcohol consumption and tobacco consumption over a 62 year period in a high alcohol consumption country: a trend analysis. BMC Research Notes 8(1):822.

Kim H-J, Fay MP, Feuer EJ, Midthune DN (2000) Permutation tests for joinpoint regression with applications to cancer rates. Statistics in Medicine 19: 335–351.

Kim H-J, Yu B, Feuer EJ (2009) Selecting the number of change-points in segmented line regression. Statistica Sinica 19(2): 597–609.

Kim J, Kim H-J (2016) Consistent model selection in segmented line regression. Journal of Statistical Planning and Inference 170: 106–116.

Lerman PM (1980) Fitting segmented regression models by grid search. Journal of Royal Statistical Society. Series C (Applied Statistics) 29(1): 77–84.

Mahapatra P (2010) An overview of Sample Registration System in India. Available at http://unstats.un.org/unsd/vitalstatkb/KnowledgebaseArticle50447.aspx

Mahapatra P (2017) The Sample Registration System in India. An overview as of 2017. Hyderabad, Institute of Health Systems.

Mari Bhat PN (1987) Mortality in India: Levels, Trends and Patterns. Pennsylvania, University of Pennsylvania.

Mari Bhat PN (2002) Completeness of India’s Sample Registration System. An assessment using the general growth balance method. Population Studies, 56(2): 119–134.

Missikpode C, Peek-Asa C, Young T, Swanton A, Leinenkugel K, Torner J (2015) Trends in non-fatal agricultural injuries requiring trauma care. Injury Epidemiology 4;2(1): 30.

Mogos MF, Salemi JL, Spooner KK, McFarlin BL, Salihu HM (2016) Differences in mortality between pregnant and nonpregnant women after cardiopulmonary resuscitation. Obstetrics and Gynecology 128(4): 880–8.

National Institute of Health (2020) Joinpoint Regression Program, Version 4.8.0.1. Bethesda, National Institute of Health, National Cancer Institute. Surveillance Research Program. Division of Cancer Control and Population Sciences.

Pollard JH (1982) The expectation of life and its relationship to mortality. Journal of Institute of Actuaries 109: 225–240.

Puzo Q, Qin P, Mehlum L (2016) Long-term trends of suicide by choice of method in Norway: a joinpoint regression analysis of data from 1969 to 2012. BMC Public Health 16:255.

Qiu D, Katanoda K, Marugame T, Sobue T (2009) A Joinpoint regression analysis of long-term trends in cancer mortality in Japan (1958–2004). International Journal of Cancer 124: 443–448.

Rea F, Pagan E, Compagnoni MM, Cantarutti A, Pigni P, Bagnardi V, Cprrap G (2017) Joinpoint regression analysis with time-on-study as time-scale. Application to three Italian population-based cohort studies. Epidemiology, Biostatistics and Public Health 14(3): e12616.

Tyczynski JE, Berkel HJ (2005) Mortality from lung cancer and tobacco smoking in Ohio (U.S.): will increasing smoking prevalence reverse current decreases in mortality? Cancer Epidemiol Biomarkers Preview. United States 14(5):1182–7.

United Nations (2004) World Population Prospects. The 2004 Revision. Volume III.

Analytical Report. New York, Department of Economic and Social Affairs. Population Division.

United Nations (2019) World Population Prospects 2019. Online Edition. Revision 1. New York, Department of Economic and Social Affairs. Population Division.

United Nations Population Fund (2004) Program of Action Adopted at the International Conference on Population and Development, Cairo 5-13 September, 1994. New York, United Nations Population Fund.

Zhang NR, Siegmund DO (2007) A modified Bayes Information Criterion with applications to the analysis of comparative genomic hybridization data. Biometrics 63: 22–32.

